# Immunity post-COVID-19 recovery boosts the antibody immune response to SARS-CoV-2 vaccination

**DOI:** 10.1101/2022.02.18.22271130

**Authors:** Fawzi Ebrahim, Salah Tabal, Yosra Lamami, Inas M Alhudiri, Salah Edin El Meshri, Samira M Al Dwigen, Ramadan Arfa, Asma Alboeshi, Hafsa A. Alemam, Fauzia Abuhtna, Rabeeah Altrhouni, Mohamed B Milad, Nada A Elgriw, Mahmoud A Ruaua, Zakarya Abusrewil, Warda Harroush, Mwada Jallul, Fouziyah S Ali, Farag Eltaib, Adam Elzaghied

## Abstract

Measurement of strength and durability of SARS-COV-2 antibody response is important to understand the waning dynamics of immune response to both vaccines and infection. The study aimed to evaluate the level of IgG antibodies against SARS-CoV-2 and their persistence in recovered, naïve and vaccinated individuals. We randomly investigate Anti-spike RBD IgG antibody responses in 10,000 individuals, both following infection with SARS-CoV-2 and immunization with SARS-COV-2 adenoviral-vector and killed vaccines. Overall, antibody titres in recovered vaccinated individuals and naïve vaccinee persists beyond 20 weeks. The mean levels of anti-spike IgG antibodies were higher in vaccinated participants with prior COVID-19 infections than in individuals without prior infection. Decline for IgG antibodies were faster in vaccinated individuals without previous COVID-19 infection compared to those with previous COVID-19 infection. Vaccination with adenoviral–vector vaccines generates higher antibody titers than for killed virus vaccine. Slightly over half of asymptomatic unvaccinated individuals develops antibody response. Previous COVID-19 infection elicited robust and sustained levels of SARS-CoV-2 antibodies in vaccinated individuals. A single dose of the vaccine is likely to provide greater protection against SARS-CoV-2 infection in individuals with prior SARS-CoV-2 infection, than in SARS-CoV-2-naive individuals. This study also underlines that asymptomatic infection equally generates antibodies as symptomatic infection. Those vaccinated with inactivated vaccine may require more frequent boosters than those vaccinated with an adenoviral vaccine. These findings are important for formulating public health vaccination strategies during COVID-19 pandemic.

**Importance:** Measurement of strength and durability of SARS-COV-2 antibody response is important to understand the waning dynamics of immune response to both vaccines and infection. We randomly surveyed 10,000 people for SARS-COV-2 antibodies. One vaccine dose with prior infection generated stronger immune response than two vaccine doses. Overall, antibody titres in recovered vaccinated individuals and naïve vaccinee persists beyond 20 weeks. Vaccination with adenoviral–vector vaccines generates higher antibody titers than for killed virus vaccine slightly over half of asymptomatic unvaccinated individuals develops antibody response. This study emphasise on the benefit of vaccination in inducing strong immune response. These findings are important for formulating public health vaccination strategies during COVID-19 pandemic.

## Introduction

SARS-CoV-2, the virus that causes COVID-19 has infected more than 304 million people around the world as of January 9, 2022, causing more than 5.4 million deaths (1). Both vaccination and herd immunity are important for controlling pandemics (2). However, the only way to reach herd immunity is by vaccination rather than by natural infection (3), which makes vaccination crucial (2). Vaccines stimulate different parts of the immune system, including humoral immunity. Therefore, production of neutralizing antibodies (IgG, IgM) is an indicator of vaccine efficacy. This is also observed after COVID-19 infection (4). SARS-CoV-2 causes either symptomatic or asymptomatic infection (5) and induces the production of neutralizing antibodies in most patients (6) within two weeks after the onset of symptoms (7). The duration of the humoral immune response and its protective efficacy are still unclear. Several studies have measured the levels of neutralizing antibodies induced by vaccination against SARS-CoV-2, which may play an important role in controlling viral infection (8). Detection of SARS-CoV-2 antibodies indicates current or past infection (9). However, the durability of antibody responses and their stability over time remain debatable. Regardless of natural infection or post vaccination. Some studies reported that antibodies did not decline within four months after infection (10). Others showed rapid decline of antibody levels, their late appearance, and even seronegativity (11). Some reports estimated that antibodies and memory cells against SARS-CoV-2 persist for at least 4–8 months ^,14^ Another study reported that IgG levels persist for at least nine months after exposure to the virus (15). So far, the immunity obtained from vaccination is not lifelong or even very long-lasting, but it seems to control the infection to some extent (2).

In Libya, since the vaccine campaign began in April 2021, four types of vaccines have been administered. According to the National Centre for Disease Control in Libya, more than one million individuals have been vaccinated with the first dose and a total 816,927 second doses have been administered (16). However, there was a considerable delay in provision of the 2^nd^ dose of Sputnik V and Sinovac which caused interruption of the vaccine schedule. Evaluation of immunity after vaccination is important for determining the protection provided by the vaccine. Herein we aimed to evaluate the levels of IgG antibodies against SARS-CoV-2 and their persistence after infection and/or post-vaccination.

## Material and methods

### Participants and study design

We investigated anti-spike IgG antibody responses following infection with SARS-CoV-2 and/or immunization with the first dose or with two doses of the vaccines in use in Libya. The study was conducted on the Libyan general population in several Northern cities. The study started with initial recruitment of 15,000 participants between August 15^th^ and December 31^st^ 2021. The participants were recruited from people visiting healthcare centers and employees in state institutions. The inclusion criterion was age > 18 years and exclusion criteria were pregnancy, blood or plasma transfusion during the three months preceding the study, immunosuppressive therapy, recent chemotherapy, autoimmune diseases, and renal dialysis. Blood samples of 3 ml were obtained by venipuncture using Vacutainer tubes. The samples were coded, and serum was separated and stored at 20°C until analyzed within 48 hours.

A self-administered questionnaire was used to collect data on sex, age, the type of vaccines received, the date(s) of vaccination, side effects, severity of symptoms, previous COVID-19 infection, and whether the infection (if there was one) was before or after receiving the vaccine. Information on past medical history and influenza vaccination status were also noted. Throughout this paper the term full vaccinations indicates two vaccine doses. Partially completed questionnaires which didn’t contain responses for vaccination status, date of vaccination and COVID-19 infection status were excluded from further analysis. In addition, surveys which fit exclusion criteria were also removed from data analysis

The Bioethics Committee at the Biotechnology Research Center in Tripoli, Libya (Ref No. BEC-BTRC 8-2020) approved the study. The study protocol was compatible with the World Medical Association Declaration of Helsinki (Ethical Principles for Medical Research Involving Human Subjects). All participants provided written informed consent to participate. Those who agreed to participate were given an information sheet detailing the study aim, pledging anonymity of their information, and explaining that they have the right to withdraw from the study at any time.

### Detection of SARS-CoV-2 specific serum antibodies

Beckman Coulter Access Anti-SARS-CoV-2 IgG assay was used on a UniCel Dxl 600 Access Immunoassay System to detect anti-SARS-CoV-2 antibodies according to the manufacturer’s instructions (Beckman Coulter, Germany. A sample was considered reactive (positive) for anti-S IgG if the result was ≥ 10 AU/ml.

### Statistical analysis

A web application was developed with PHP, MySQL and JavaScript specifically for electronically collecting survey data and initial statistical analysis. However, the statistical analysis was performed using Microsoft Excel and GraphPad Prism version 9.3. The descriptive statistics included mean, standard deviation and percentages. The differences between mean values were compared by the unpaired Student’s t-test. P values < 0.05 were considered statistically significant.

## Results

### Participant selection and characteristics of study group

Between August 15^th^ and December 31^st^ 2021, 10,000 adults were enrolled to the study, including both symptomatic and asymptomatic COVID-19 recoveries, as well as persons vaccinated with the first dose or with both doses of the vaccine (Figure 1).

**Fig. 1.**
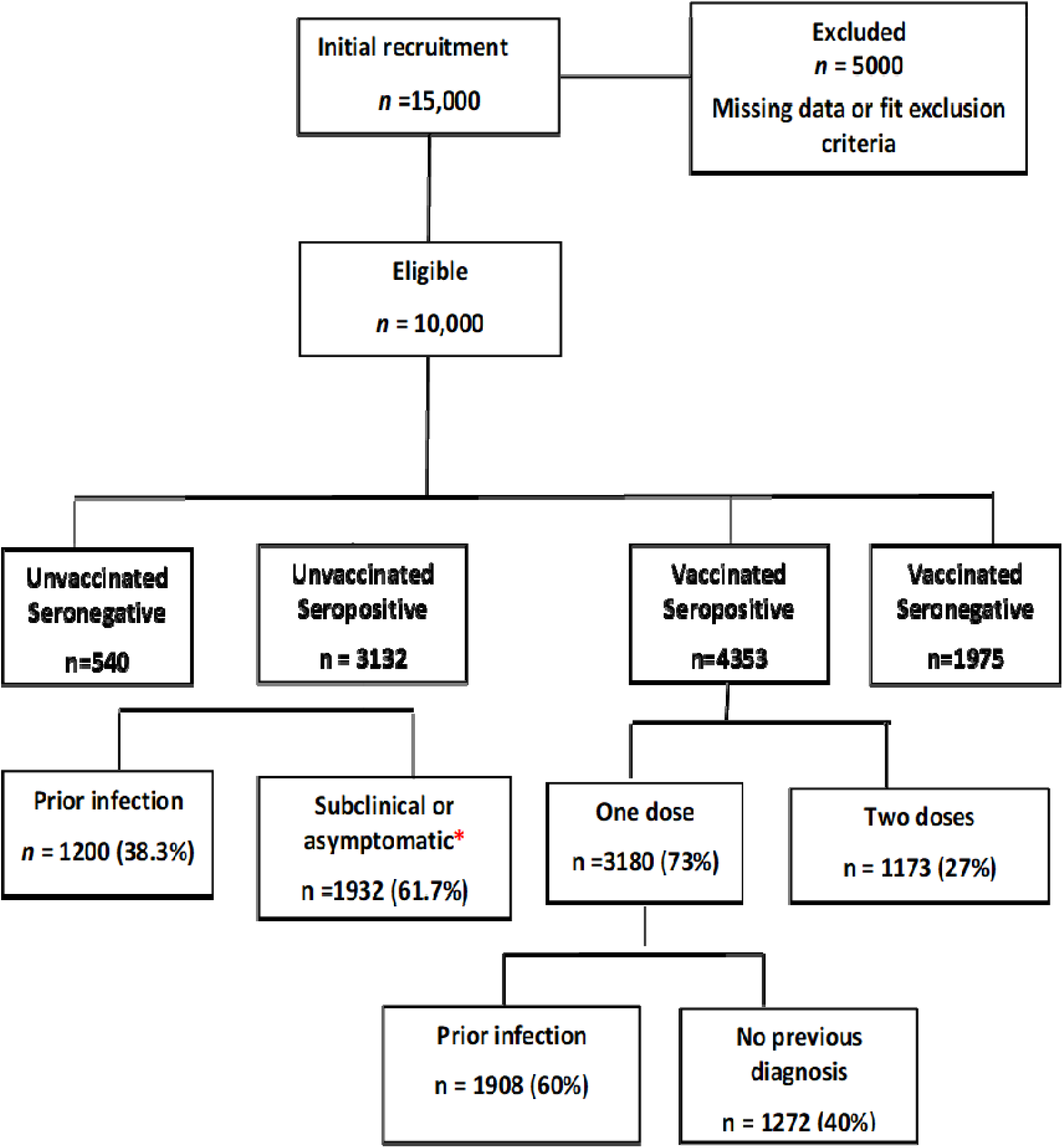
Flowchart showing recruitment and participant selection. **\***Denotes participants who didn’t report previous infection in the survey but gave positive IgG titer indicating previous undiagnosed SARS-COV-2 infection.

### Overall view of vaccinations and seropositivity

Of all the participants, 63.3% (6328/10,000) had been vaccinated. Among the 3132 unvaccinated individuals who were seropositive, 1932 did not report a previous infection, pointing to a rate of asymptomatic COVID-19 of 61.7%. The most frequently used vaccines were Sputnik V (n = 4156, 65.7%) followed by AstraZeneca (n = 1065, 16.7%) and collectively 17.5% (579/6328) of the participants had been vaccinated with either Sinopharm or Sinovac (Table 1). The overall rate of positivity was lowest for the Sinovac and Sinopharm vaccines (52.2% and 52.4%, respectively) and considerably higher for the AstraZeneca and Sputnik V vaccines (73.8% and 71.9%) (Table 1).

**Table 1.** Distribution of vaccinated participants by sex, vaccine type, and IgG seropositivity.

| Vaccine | n |  |  | Seropositive |  | Seronegative |  |
| --- | --- | --- | --- | --- | --- | --- | --- |
|  | Females | Males | Total | n | % | n | % |
| Sputnik V | 1874 | 2282 | 4156 | 2988 | 71.9 | 1168 | 28.1 |
| AstraZeneca | 479 | 586 | 1065 | 786 | 73.8 | 279 | 26.2 |
| Sinopharm | 237 | 257 | 494 | 259 | 52.4 | 235 | 47.6 |
| Sinovac | 306 | 307 | 613 | 320 | 52.2 | 293 | 47.8 |
| All vaccines | 2896 | 3432 | 6328 | 4353 | 68.8 | 1975 | 31.2 |

### Analysis of seropositivity by age, vaccination and prior infection status

The distribution of seropositivity rates by age group, prior infection status and vaccination status is shown in table 2. Notably, all participants who had been administered one dose were seropositive, with the exception of those below 35 years of age and not previously infected, the seropositivity rate was 87.9% (Table 2). An incline in positivity rate in the eldest age group was also noticed in fully vaccinated individuals regardless of infection status (Table 2).

**Table 2.** Seropositive individuals by age group, vaccination status and prior infection status.

| | $\leq 35$ years | | 36–45 years | | 46–55 years | | 56–65 years | | > 65 years | |
| --- | --- | --- | --- | --- | --- | --- | --- | --- | --- | --- |
|  | p =0.0001 |  | p<0.0001 |  | p=0.0003 |  | p<0.0001 |  | p=0.0002 |  |
|  | *N* | % | *N* | % | *N* | % | *N* | % | *N* | % |
| **Unvaccinated** |  |  |  |  |  |  |  |  |  |  |
| Prior infection | 386 | 77.0 | 320 | 96.0 | 307 | 93.0 | 147 | 95.0 | 40 | 88.8 |
| No prior infection | 597 | 62.8 | 484 | 74.3 | 536 | 82.0 | 206 | 92.0 | 102 | 78.0 |
| **Vaccinated with one dose** |  |  |  |  |  |  |  |  |  |  |
| Prior infection | 358 | 100 | 582 | 100 | 419 | 100 | 308 | 100 | 383 | 100 |
| No prior infection | 585 | 87.9 | 637 | 100 | 678 | 100 | 443 | 100 | 354 | 100 |
| **Vaccinated with two doses** |  |  |  |  |  |  |  |  |  |  |
| Prior infection | 150 | 100 | 149 | 94.2 | 164 | 100 | 56 | 97.0 | 227 | 93.8 |
| No prior infection | 210 | 72.8 | 129 | 79.3 | 184 | 82.8 | 126 | 86.9 | 188 | 90.9 |

Males represented 54.2% of the study sample. The ages of the participants ranged from 18 to 90 years. The majority of participant were aged between 36-45 years. Antibody titers against spike antigen were similar in both sexes in all vaccines except for Sinovac which showed significantly higher titers in female participants (p=0.026). However, the highest median titers were detected in individuals who received AstraZeneca vaccine (Fig.2). In females >65 years who received a single dose of Sinovac with no previous infection the mean IgG titer was 151 AU/ml compared to 48.77 AU/ml in males.

Overall, the seropositivity rate was higher among unvaccinated individuals who reported prior infection (convalescent) than those who did not (asymptomatic or subclinical) (Table 2). In unvaccinated convalescent individuals, seropositivity was relatively similar in the five age groups, ranging from 77% (≤ 35 years) to 96% (36-45 years). In contrast, seropositivity among unvaccinated individuals with no prior infection increased with age from 62.8% (< 35 years) to 92% (55-65 years), after which there was a dip to 78% (Table 2).

### Waning dynamics of IgG anti-spike RBD levels

From week 1 after single dose vaccination in recovered participants, the mean titers of IgG against the spike protein increased for all vaccines but were considerably higher for the AstraZeneca and Sputnik V vaccines, which, by week 11, reached 169.2 and 142.0 AU/ml, than for Sinopharm and Sinovac (92.4 and 99.3 AU/ml). However, while the levels against Sputnik V and AstraZeneca peaked on week 7 and then started a gradual decline, the IgG levels against Sinovac and Sinopharm continued to rise until week 15, when the levels against the four vaccines became very similar, ranging from 113.6 AU/ml (Sinopharm) to 123.8 AU/m (Sputnik V) (Figure 2). Thereafter, the rate of decline was more notable for AstraZeneca and Sinopharm. From week 19 to week 21, anti-Sputnik V and anti-Sinovac IgG levels were the highest.

**Fig. 2.**
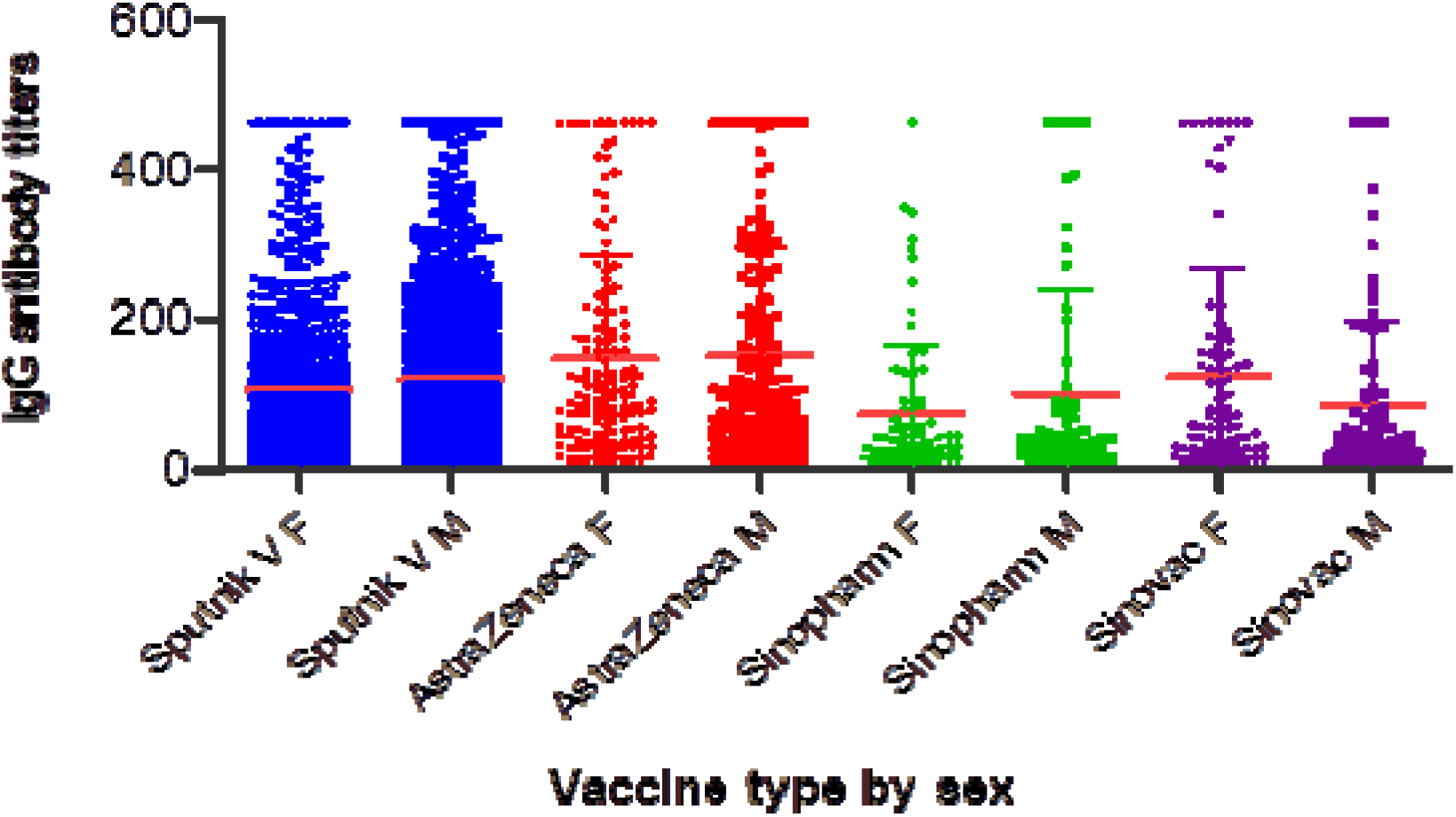
Anti-spike IgG titers developed after each vaccine type by sex. F denotes female and M denotes male. The red line represents the median IgG antibody titers for each group. The chart shows similar median IgG titre in both male and females except in the group who received sinovac vaccine where females had significantly higher antibody response than males.

In seropositive participants without evidence of prior infection, post-vaccine antibody responses showed that positive anti-spike IgG results increased over the 3–4 weeks after the first vaccination in case of Sputnik V, AstraZeneca and Sinovac respectively (Figure 3). In terms of Sinopharm, the titration elevated over the 5th week. The higher average of titration IgG was observed in Sputnik V in week 12 (108.36 AU/ml. P value <0.005) while in case of AstraZeneca the maximum antibody titration was seen in week 10 (132.67 AU/ml. P < 0.005). The higher titration of seropositive value were (93.7 and 91.05 AU/ml. P < 0.005) in Sinopharm and Sinovac respectively.

**Fig. 3.**
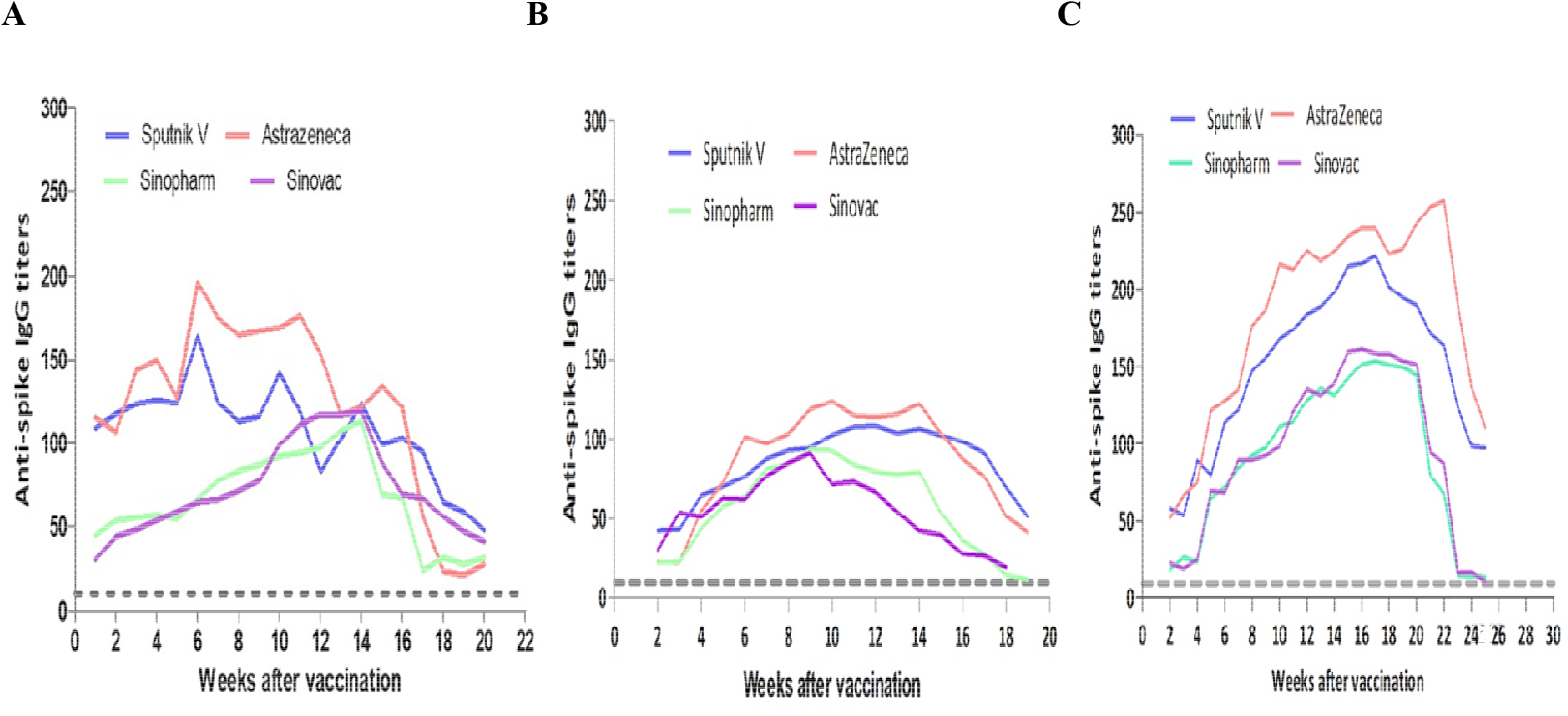
Individuals with previous SARS-COV-2 infection elicited faster and stronger antibody titers to all types of vaccinations than individuals with no previous infection (naïve). The charts shows waning dynamics in different groups according to previous infection status and vaccine dose. **A**. Average anti-spike IgG titers after first vaccination dose in previously infected participants. **B**. Average anti-spike IgG titers after first vaccination dose in seropositive participants who never had COVID-19 symptoms. **C** Trajectory of average of anti-spike IgG titers in fully vaccinated individuals (two doses). The grey dashed line in each graph indicates the threshold antibody positivity at 10 AU/ml.

In fully vaccinated respondents (two doses), the average titers of anti-spike IgG in participants who had received AstraZenca or Spunik vaccine were 257 AU/ml and 221 AU/ml, respectively (p value < 0.005), which is considerably higher than in those who had received only the first dose with or without prior infection (Figure 4). Similar observation were seen in Sinopharm and Sinovac as compared with patients who received the first dose (figures 2 and 3).

## Discussion

The persistence and robustness of antibody response to vaccination and infection are important parameters for assessment of vaccine efficacy and natural immune response. Public health administrations require evidence for the need of booster doses and which vaccines perform better than the others. In addition, because of mostly poor compliance with protection measures, a great proportion of young population, and reduced COVID-19 testing in Libya, estimation of natural induced immunity was important to evaluate the exact epidemiological burden of the disease. Our study sought to answer these questions through analysis of anti-spike RBD antibodies in a random population of 10,000 individuals.

The study showed that nearly two-thirds of the unvaccinated seropositive had been exposed to the virus and were asymptomatic but were not previously diagnosed. This highlights that both symptomatic and asymptomatic or subclinical SARS-COV-2 infection generates good immune response. Furthermore, this prevalence in the community indicates the widespread of COVID-19 and potential protection against reinfection, severe disease and hospitalization (17).

Administration of one vaccine dose in recovered individuals generated stronger immune response than two vaccine doses. Our results confirm previous findings that vaccinated individuals with prior infection is associated with higher titration of SARS-CoV-2-specific IgG antibodies than vaccinated naïve individuals (18–20). Whether these persons require a second vaccine dose is uncertain because the protective antibody level is unclear. Further studies should be performed to establish a protection threshold versus positivity threshold (21).

We also found that a single-dose of either “Sputnik V” or AstraZeneca vaccine formed a faster humoral immune response in participants who had previous infection which is similar to findings in other studies (22, 23). IgG anti-spike antibodies was triggered in less than 2 weeks particularly in patients who received first dose with prior infection in all vaccines type involved in our study. The first dose, given to individuals whose immune systems previously stimulated by the natural infection, probably had a similar effect as the second dose in naïve individuals who received the first vaccine dose. In light with the increased demand and shortage of vaccine supply, these findings would suggest implementing a strategy of a single vaccine dose for individuals who were previously infected by SARS-CoV-2.

Since the manufacturing of COVID-19 vaccines, there was questions over which vaccine performs better than the other in terms of durability and robustness. Our analysis showed that vaccination with adenoviral–vector vaccines (Sputnik V and AstraZeneca) generated higher antibody titres than for inactivated vaccines (Sinopharm and Sinovac). This finding stand in agreement with previous studies on reduced antibody titres to Sinovac and Sinopharm compared with AstraZeneca and mRNA vaccines (24–26). These results suggest that giving a third booster dose would be beneficial to prevent COVID-19 infection in inactivated vaccine vaccinee. These preliminary data serve as a basis for further studies on the correlation of incidence of breakthrough infections with declining neutralizing antibody or anti-spike IgG titers to guide booster shot administration. Many countries have offered booster doses to those who received Sinopharm and Sinovac regardless of the date of receiving the 2^nd^ dose (27).

Overall, antibody titres in recovered vaccinated individuals and naïve vaccinee persisted beyond 20 weeks for single dose recipients and beyond 24 weeks for two dose vaccine recipients. Notably, fully-vaccinated respondents with or without previous COVID-19 infection showed high titration for all vaccines but significantly higher for AstraZeneca and Sputnik V vaccines (239 AU/ml and 221 AU/ml at week 17 respectively). The decline in IgG levels over time was expected and occurred in all vaccine brands in the current study. The serological analysis in this study was performed at an average of five months post vaccination. We didn’t follow up persons beyond that period. This finding agreed with previous published data showing that antibodies detected for several months after vaccination (28). Additionally, in vaccinated individuals, IgG levels were maintained for longer periods in those who were previously infected than naïve persons (29, 30).

The current study has some limitations. The study did not include children and adolescents thus missing the actual frequency of infected population in Libya. In order to identify persons with unknown previous infection in vaccinated individuals, anti-nucleocapsid antibodies should be measured which was not the case in this study. According to the frequency estimate in the unvaccinated group, we believe that good proportion of full dose vaccinee to be previously asymptomatically infected. Furthermore, most fully vaccinated participants did not receive the second dose on schedule because of the delay in provision of vaccines in Libya, due to global shortage, which could reflect on interpretation of results in this group. Although this is not the normal scenario, but it is clear from the study that this delay didn’t affect the induction of immune response which has implications for dosing interval and vaccine administration policy.

## Conclusion

SARS-CoV-2 anti-spike IgG levels among unvaccinated individuals with previous infection either symptomatic or asymptomatic showed good immune response. Vaccination in individuals with prior COVID-19 infection demonstrated higher IgG titration than vaccination without prior infection. Anti-spike serum IgG following vaccination persisted beyond 20 weeks indicating probable protection against infection. Our results suggest that even greater efforts should be made to immunize more individuals either primarily or via booster doses, particularly people without prior COVID-19 infection. However, further investigations are needed to estimate the durability of these immune responses and the potential need for additional booster doses as antibody levels decline with time as well as antibody threshold for protection. Those vaccinated with inactivated vaccine may require more frequent boosters than those vaccinated with an adenoviral vaccine. These findings are important for formulating public health vaccination strategies during COVID-19 pandemic.

## Data Availability

All data produced in the present study are available upon reasonable request to the authors

## Author contributions

Study Conception and design: AE, FE, IA and ZA. YL, SD, WH, HE, NG, FM, FS, RT, ME, AA and MA performed the antibody titer tests. RA, ST and FE analyzed the data. MJ and FE wrote the introduction, YA wrote the methods, FE wrote the results and discussion. AZ, FE IA finalized the manuscript. All authors had full access to all the data in the study, reviewed the final manuscript, and approved its submission for publication.

## Acknowledgements

The authors thank the Ministry of Health and the Libyan Authority for Scientific Research. We also thank all the health care workers for their valuable work in blood sampling. In addition, we thank the study participants for their support of COVID-19 research.

## Conflict of interest

The authors declare that they have no conflict of interest.

## Notes

### Competing Interest Statement

The authors have declared no competing interest.

### Funding Statement

This study was funded by Ministry of Health Libya.

### Author Declarations

The study was approved by "The Bioethics Committee at the Biotechnology Research Center in Tripoli, Libya (Ref No. approved the study BEC-BTRC 8-2020).”

